# The Parkinson’s Disease Associated BAP1/ASXL3 Complex Regulates the Internalization of α-Synuclein Fibrils by Reprogramming the Cell Surface Glycoproteome

**DOI:** 10.64898/2026.09.09.26362627

**Authors:** Nathan C. Karpilovsky, Thomas Goiran, Konstantin Senkevich, Ghislaine Deyab, Nicholas Twells, Emily NaYoung Cha, Emma Macdougall, Zhangjie Wang, Julien Sirois, Graham Macleod, Lang Liu, Morvarid Ghamgosar Shahkhali, Kalidou Ali Boubacar, Jace Jones-Tabah, Isabella Pietrantonio, Wen Luo, Irina Shlaifer, Esther Del Cid-Pellitero, Ayodeji Kulepa, El Bachir Affar, Stephane Angers, Jian Liu, Lara K. Mahal, Thomas M. Durcan, Ziv Gan-Or, Edward A. Fon

## Abstract

A central hallmark of Parkinson’s disease (PD) is the spread of α-Synuclein (αSyn) aggregates, which is thought to contribute to its progressive nature. To better understand the mechanism of cellular internalization of αSyn fibrils, we conducted a genome wide CRISPR activation (CRISPRa) screen to identify genetic modifiers of fibril uptake. We report here BRCA-associated protein 1 (*BAP1*) as a regulator of fibril uptake into cells and validate its association with the genetic risk of PD. BAP1 regulates fibril entry into cells by acting as a master transcriptional regulator of the cell surface glycoproteome. BAP1 downregulates Heparan Sulfate Proteoglycan (HSPG) and upregulates O-linked glycoprotein expression, the net effect of which is to reduce fibril internalization. The effect of enhancing BAP1 expression is cell-type specific, as it reduces αSyn uptake in iPSC-derived dopaminergic neurons, but not in microglia. Using human midbrain organoids with integrated microglia, we find this effect is mediated through ASXL3, another PD risk gene and a component of the BAP1 Polycomb Repressive-Deubiquitinase Complex (PR-DUB). These findings are consistent with higher *BAP1*/*ASXL3* co-expression in regions of the brain less vulnerable to PD pathology. Together, the work uncovers a novel role for BAP1/ASXL3 in regulating αSyn fibril internalization by remodelling the cell surface glycoproteome.

## Introduction

Parkinson’s disease (PD) is a devastating neurodegenerative disorder characterized by the loss of dopamine neurons (DNs) in the substantia nigra (SN) pars compacta of the midbrain^1^. The most recognizable pathological hallmark of PD is the presence of Lewy bodies in neurons, which are evident in most PD patients^1, 2^. The major component of these pathological inclusions is aggregated forms of the protein α-Synuclein (αSyn), in addition to other misfolded proteins, organelles, and membrane-bound structures^2, 3^. Mutations in the protein coding sequence^4–6^, copy number variants (CNVs)^7–9^ and single nucleotide polymorphisms (SNPs)^10–12^ in *SNCA*, the gene encoding αSyn, have been identified to cause PD in families and increase the risk of sporadic PD in Genome-Wide association studies (GWAS). Despite its unfolded structure in solution, or a partial α helical structure upon membrane association, αSyn has a propensity to misfold^13–15^. Misfolded αSyn can aggregate into β-sheet rich fibrils, which have been shown to spread from one cell to another, where they can recruit endogenous native αSyn monomers and, in turn, induce them to misfold and aggregate in a prion-like fashion^16, 17^. This spreading of aggregation and propagation of αSyn pathology across the brain is believed to explain the progression of symptoms in patients and represents a promising target to slow disease progression^18^.

A key step in the spread of αSyn pathology is the internalization of aggregates into cells, a process that has been suggested to be regulated by several pathways, including uptake by specific receptors such as LAG-3^19^, LRP1^20^, GPNMB^21, 22^, and FAM171A2^23^, as well as both clathrin-mediated endocytosis^24^ and ultrarapid micropinocytosis^25^. The internalization of αSyn fibrils and other protein aggregates also involve heparan sulfate proteoglycans (HSPGs), a type of proteoglycan with negatively charged polysaccharide chains, which have been shown to mediate the binding of various cargo (e.g. virus binding and other proteinaceous cargo) to cellular membranes^26–28^. Heparan sulfate (HS) chains are polymorphic in length and sulfation pattern which can modulate the binding to specific cargo such as αSyn, Tau, and Aβ fibrils^29^. Despite the importance of HSPGs in the internalization of αSyn fibrils, there is still much that remains to be discovered about their role, mechanisms, and regulation in synucleinopathies. Moreover, almost nothing is known about whether other types of glycosylation affect αSyn fibril internalization into cells.

Here, using a genome-wide CRISPR activation (CRISPRa) screen, we identify the deubiquitinase BRCA-associated protein 1 (*BAP1*) as a novel genetic regulator of αSyn fibril internalization. BAP1 is a component of the Polycomb Repressive-Deubiquitinase Complex (PR-DUB) complex involved in transcriptional regulation of target genes by controlling histone H2AK119 mono-ubiquitination (H2AK119Ub)^30^. *BAP1* was previously mapped to a PD GWAS locus^11^ but this was not replicated in subsequent GWAS’^10, 12^. Here, using a combination of fine mapping and brain wide expression data of variants in the *BAP1* locus combined with Mendelian randomization, we show that *BAP1* expression affects PD risk. Increased BAP1 expression lowers PD risk by transcriptionally reprogramming the cell surface glycoproteome. In addition to reducing the levels of HSPGs at the cell surface, BAP1 expression increases O-linked mucin-type protein glycosylation at the plasma membrane, which in combination with the reduction in HSPGs, reduces αSyn fibril binding to the cell surface. These effects of BAP1 are cell-type specific, affecting αSyn fibril internalization in DNs but not in microglia. Mechanistically, this specificity is likely mediated by ASXL3, another PD risk gene, which is also a tissue-specific component of the PR-DUB complex^31–33^. Indeed, using single cell RNA sequencing in a midbrain organoid model of PD containing integrated microglia, we show that *ASXL3* is not expressed in microglia compared to DN and other neuronal populations^34^. Moreover, we show that, in aged organoids, the expression of both *BAP1* and *ASXL3* are reduced in SN-like (SOX6+) DNs, which are most vulnerable to degeneration in PD^35, 36^, compared to ventral tegmental area-like (SOX6-) DNs, which are more resilient and typically spared early in the course of PD. Together, our work implicates two understudied PD risk genes that function in the PR-DUB complex to regulate the internalization of αSyn fibril and DN vulnerability by transcriptionally reprogramming the cell surface glycoproteome.

## Results

### CRISPRa screen nominates *BAP1* as a modifier of αSyn fibril uptake

To identify genes that modulate uptake of αSyn fibrils into cells, we used retinal pigment epithelium cells (RPE1) stably expressing dCas9-VPR^37^ transduced with a genome-wide sgRNA library (hCRISPRa-v2)^38^, to screen for the internalization of Alexa 488 labelled αSyn preformed fibrils (PFFs) (fig 1a-b and supplemental fig 1a-d). PFF-treated cells were sorted by FACS into groups exhibiting high- and low-fluorescence, corresponding to populations of cells with high and low PFF internalization, respectively (supplemental fig 1e), revealing significantly over- and under- represented genes^39^ (fig 1c-d and supplemental table 1). To determine whether the screen identified previously reported pathways, Over Representation Analysis (ORA) on the top 100 positively associated and top 100 negatively associated genes was performed through WebGestalt^40, 41^ (fig 1e). The top enriched term was identified as *glycosaminoglycan biosynthetic process*, in agreement with previous literature describing HSPGs to be involved in the internalization of αSyn into cells^26–29^. To identify genes from the screen that have been previously linked genetically to PD, we cross-referenced the top 50 positively associated and 50 negatively associated genes with those nominated in PD GWAS (2017, 2019, and 2025) (fig 1f), revealing *BAP1*, as a potential PD risk gene involved in regulating αSyn fibril uptake, prompting us to study it further.

**Figure 1:**
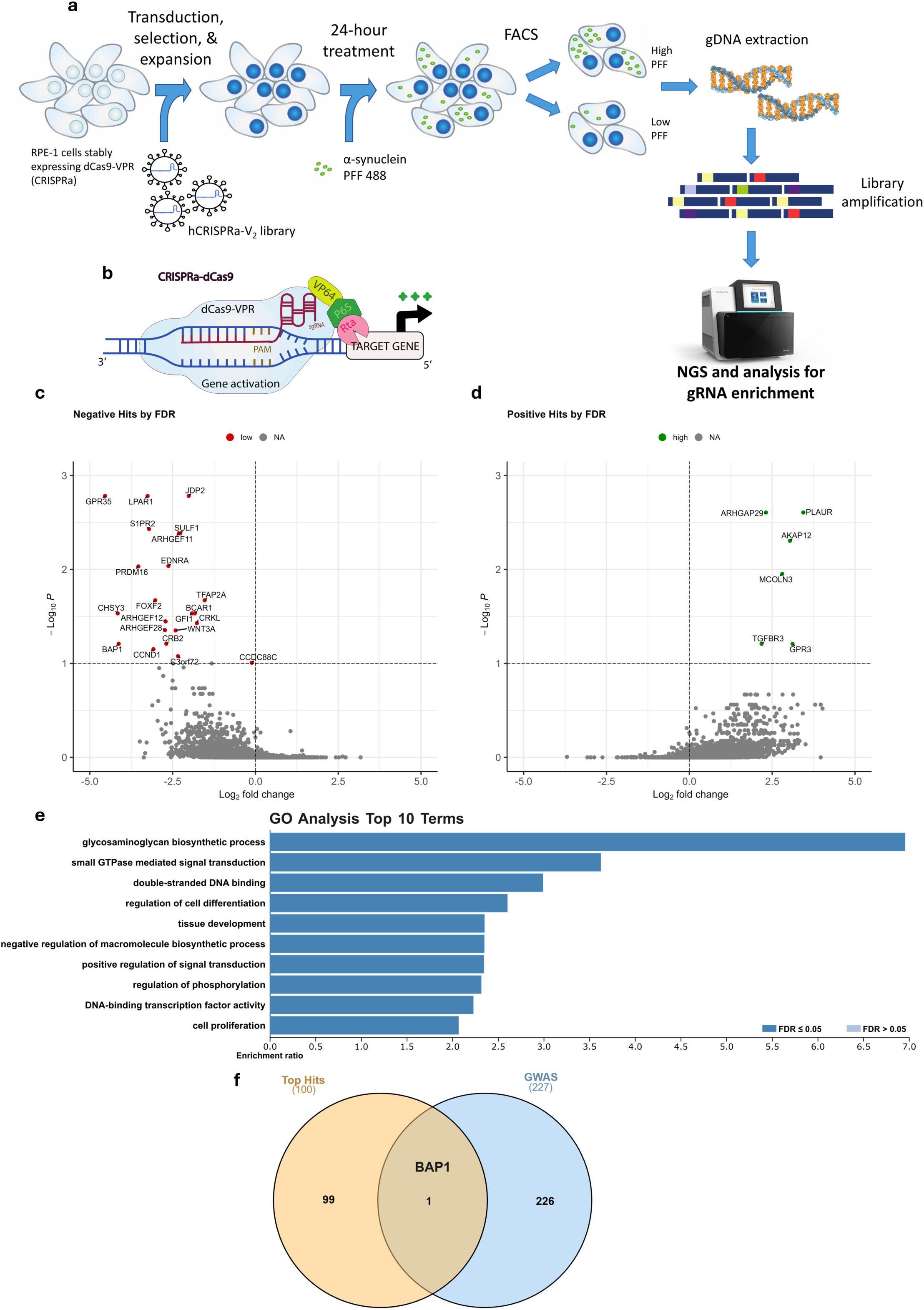
CRISPRa screen reveals *BAP1* as a negative modifier of αSyn fibril uptake. a) CRISPRa screen schematic and workflow. b) CRISPRa, dCas9-VPR, machinery schematic used in the CRISPRa screen. c) Volcano plot of negative modifiers of PFF uptake through MAGeCK^39^ analysis. Genes that potentially decrease αSyn fibril uptake are marked in red. d) Volcano plot of positive modifiers of PFF uptake through MAGeCK^39^ analysis. Genes that potentially increase αSyn fibril uptake are marked in green. e) Over representation analysis (ORA) of 100 positive and 100 negative modifiers of PFF uptake from the CRISPRa screen, using WebGestalt with weighted set reduction^40, 41^. f) Overlap between top 100 bidirectional modifiers of PFF uptake from CRISPRa screen with GWAS-nominated PD risk genes, showing *BAP1* as the only overlapping gene.

### Brain expression analysis and Mendelian randomization implicate *BAP1* as a PD risk gene

*BAP1* was nominated within a PD risk locus in the 2017 GWAS^11^. However, this locus did not reach genome-wide significance in subsequent GWAS’^10, 12^. To determine whether *BAP1* could mediate PD risk, we examined *BAP1* expression in brain using GTEX. Notably, *BAP1* expression is highest in regions least vulnerable to degeneration in PD such as the cortex and cerebellum, whereas it is lowest in vulnerable regions such as the SN and basal ganglia (fig. 2a), consistent with Keo and colleagues^42^, showing that *BAP1* expression is inversely correlated with vulnerability and disease progression in the Braak staging system^43, 44^. Next, we explored whether genetic variants in the *BAP1* locus associated with PD risk influence the levels of *BAP1* expression in brain tissues. The variant rs123598 showed strong association with both PD risk (GWAS) and *BAP1* expression (eQTL), suggesting that the same genetic variant may be influencing both disease risk and gene expression (fig. 2b). When also considering the variant rs34332947, which is in linkage disequilibrium with rs123598 (D′ = 1, r² = 1; fig 2b-c), we observe a consistent reduction in *BAP1* expression associated with the risk alleles of both SNPs compared to their respective control alleles in Nucleus Accumbens, Caudate, Hippocampus, and Cortex (fig 2c, e-f). This suggests that decreased *BAP1* expression is associated with increased risk of PD. To determine whether the link between PD risk and *BAP1* expression is causal, we utilized Summary data based Mendelian Randomization (SMR). The PD risk allele increases disease risk (β_GWAS = 0.054, P = 5.4 × 10⁻⁴) while reducing *BAP1* expression (β_eQTL = −0.478, P = 7.3 × 10⁻¹²), indicating that decreased *BAP1* expression mediates genetic risk (fig 2d). SMR confirmed this relationship, with a negative SMR effect (β_SMR = −0.114, P = 2.0 × 10⁻³) consistent with genetically predicted higher *BAP1* expression being associated with lower PD risk. Colocalization analysis of the GP2 PD GWAS with brain cortex eQTLs further nominated *BAP1* (PP.H4 = 0.58) and *STAB1* (PP.H4 = 0.68) as candidate effector genes at this locus. Although neither exceeded the conventional threshold for strong colocalization (PP.H4 > 0.8), the concordant GWAS–eQTL overlap together with the SMR provide additional genetic support for *BAP1* as a candidate effector gene at this locus. Taken together, these results point to the potential causal involvement of *BAP1* in PD, where *BAP1* expression is inversely associated with PD risk.

**Figure 2:**
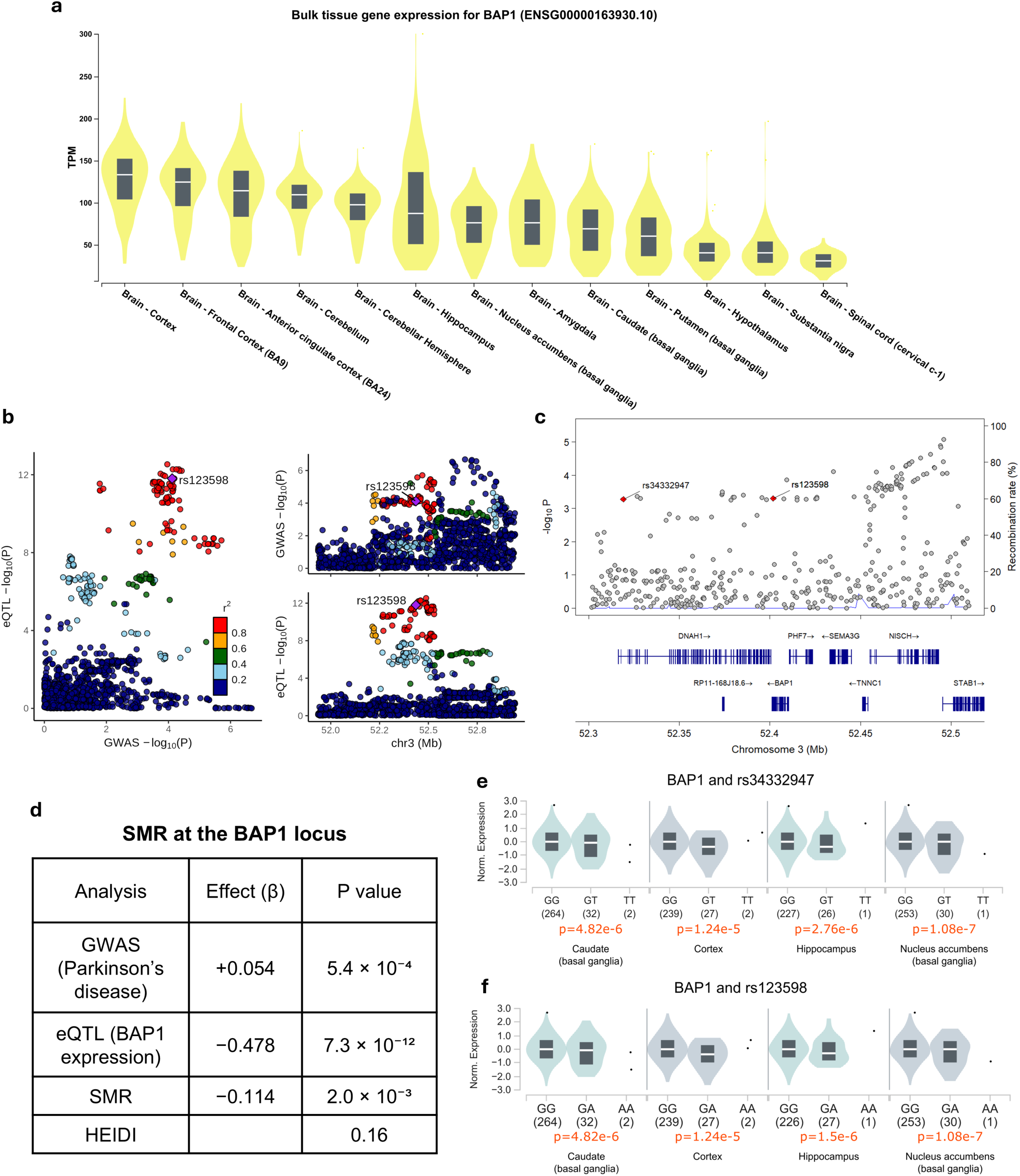
Genetic analysis of *BAP1* expression and association to PD risk. a) GTEX bulk tissue expression for *BAP1* across brain regions. b) LocusZoom plot comparing the *BAP1* locus (±100 kb) in Parkinson’s disease GWAS and brain eQTL meta-analysis. Parkinson’s disease GWAS association is plotted together with brain eQTL association across the region, with variants colored by linkage disequilibrium (r²) to the lead SNP rs3432947, allowing visual assessment of shared regulatory and disease signals. c) Regional association and recombination landscape at the *BAP1* locus. LocusZoom plot displays Parkinson’s disease GWAS association across the *BAP1* region, with rs3432947 and rs123598 highlighted. Both variants lie within the same association peak and are in complete linkage disequilibrium (r^2^=1 and D’=1), representing the same regulatory haplotype at this locus. d) Summary-based Mendelian randomization results for *BAP1*. The table reports GWAS, eQTL, and SMR effect estimates for the lead variant (rs3432947). HEIDI testing shows no evidence of heterogeneity (P = 0.16). The Parkinson’s disease risk allele increases disease risk while reducing *BAP1* expression. The negative SMR effect therefore indicates that higher *BAP1* expression is protective. e) eQTL normalized expression levels correlated with *BAP1* rs34332947 SNP in brain regions. Each brain region, including caudate, cortex, hippocampus, and nucleus accumbens. P-values shown under each graph along with sample size, showing decreased levels of *BAP1*. f) eQTL normalized expression levels correlated with *BAP1* rs123598 SNP in brain regions. Each brain region, including caudate, cortex, hippocampus, and nucleus accumbens. P-values shown under each graph along with sample size, showing decreased levels of *BAP1*.

### BAP1 levels regulate the accumulation of αSyn fibrils in cells

Based on the results of our screen and the genetic evidence above, we next asked whether BAP1 regulates the accumulation of αSyn fibrils in cells and the mechanisms involved. Using lentivirus-delivered sgRNAs against *BAP1* into RPE1 cells stably expressing the CRISPRa dCas9 machinery, we found that enhancing BAP1 expression (fig 3a-c), resulted in a substantial reduction of αSyn PFF accumulation by flow cytometry (fig 3d-f) and high content microscopy over 24 hours (fig 3g-h). Since processing cells for flow cytometry involves removal of residual αSyn PFF at external face of the cell surface by trypsinization, our results indicate that BAP1 activation reduces PFF internalization.

**Figure 3:**
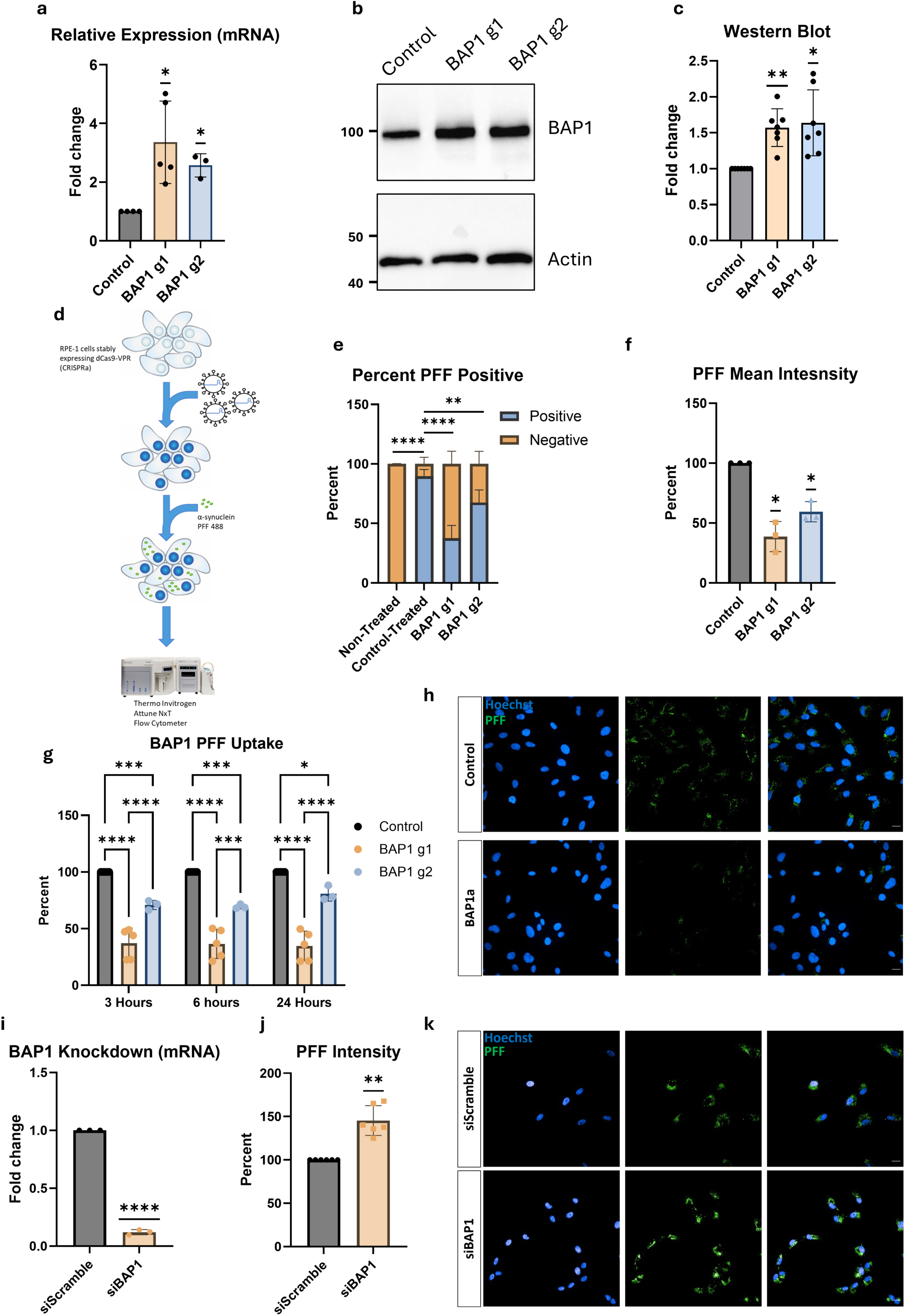
BAP1 bi-directionally regulates internalization of αSyn fibrils in cells. a) RT-qPCR of *BAP1* levels comparing BAP1a g1 and BAP1a g2 to control cells, to validate overexpression. n=3-4 biological replicates. b) Representative western blot against BAP1 (top) and Actin (bottom, loading control) comparing BAP1a g1 and BAP1a g2 to control cells. c) Quantification (densiometric analysis) of western blots with ImageJ, n=7 biological replicates. d) Layout of flow cytometry experiments to measure 3 hour PFF uptake in BAP1a cells. e) Percent PFF positive/negative cells in each population. Gated using non-treated cells as negative control, each population is graphed as a percentage of PFF positive and negative cells. n=3 biological replicates. f) Mean intensity measurements of PFF 488 fluorescence in BAP1a cells compared to control. Each biological replicate normalized to its baseline control. n=3 biological replicates. g) High content confocal microscopy PFF uptake assay at 3, 6, and 24 hours. After background subtraction BAP1a g1 was normalized to its control at each time point. n=5 biological replicates. h) Representative image of BAP1a g1 vs control CRISPRa cells at 3 hours post PFF treatment. Hoechst (blue) and PFF (green). Scale bar 20 µm. i) RT-qPCR of *BAP1* siRNA-mediated knockdown compared to siScramble. n=3 biological replicates. j) 24 hour PFF uptake assay on BAP1 siRNA-knockdown, normalized to control within each experiment. n=6 biological replicates. k) Representative image of PFF uptake assay showing increased PFF signal (green) in BAP1 knockdown compared to siScramble. Hoechst (blue) and PFF (green). Scale bar 20 µm. Statistical Analysis: a) two-way ANOVA with Šídák’s multiple comparisons test. c,e,h,i,j) one sample t-test. g) 2-way ANOVA with Dunnett’s multiple comparisons test. Graphs are presented as mean ± SD. *p-value<0.05, **p-value<0.01, *** p-value<0.001, ****p-value<0.001

We also treated BAP1 CRISPRa cells with fluorescently labelled cargo, including 10k Dextran, EGF, transferrin, and Tau oligomers, to identify whether the effect of BAP1 is specific to αSyn fibrils or affect general internalization pathways. BAP1 CRISPRa cells showed no statistically significant difference with control cells in the internalization of the tested fluorescent cargo by high content microscopy, suggesting that the effect is specific to αSyn fibrils (supplemental fig 2a-d). Moreover, we utilized a pH-sensitive (pHrodo) to label PFFs to ensure that fibrils are internalized into lysosomal compartments as previously reported^25, 37^. As expected, we find that, compared to the control cells, the decrease in internalization of αSyn fibrils in BAP1 CRISPRa cells leads to a decrease in their accumulation in acidic endolysosomal compartments over time (supplemental fig 2e). We then sought to determine whether conversely, a reduction of BAP1 could increase αSyn fibril uptake, in line with PD risk. Using siRNA-mediated knockdown, we effectively reduced BAP1 expression, which led to increased PFF uptake into RPE1 cells compared to scrambled siRNA transfected control cells (fig 3i-k). These results suggest that BAP1 expression bi-directionally regulates αSyn fibril uptake, congruently with the direction of genetic risk.

### BAP1 activation reduces cell surface HSPG levels and αSyn fibril binding

As HSPGs have been shown to play role in αSyn fibril internalization by promoting cell surface binding of PFFs^27, 28^, we asked whether HSPGs also mediated the effect of BAP1 on αSyn fibril internalization. Using an assay to concurrently measure cell surface PFF-binding and HSPG levels by incubating cells at 4 °C (to arrest membrane trafficking and block internalization) with an anti-HSPG antibody and with Alexa 488 labelled PFFs (fig 4a), we observed that BAP1 activation leads to a significant reduction in both cell surface HSPG levels and PFF binding (fig 4b-d). To confirm BAP1’s role in regulating the glycoproteome and uncover whether there are specific HS chain differences, we performed a disaccharide analysis via mass spectrometry. Trypsinization of cells leads to cleavage of HSPG-modified peptides exposed on the extracellular face of the plasma membrane. The cleaved HSPGs were collected from the conditioned media (cell surface fraction) and analysed by LC-MS for disaccharide content and compared to the HSPGs remaining in the cellular pellet (intracellular fraction) (fig 4e-f). The disaccharide analysis revealed a global reduction in total HSPG chains in both the cell surface and intracellular fractions. We detected no significant changes in any particular disaccharide unit, but rather a consistent reduction in all disaccharides that were analysed. Notably, △UA-GlcNAc subunits, the building block and most abundant disaccharide on HSPGs, were significantly reduced intracellularly (fig 4f). In contrast, we detected no significant differences between BAP1a and control cells in chondroitin sulfate proteoglycans (CSPGs) (fig 4g), a distinct but related class of proteoglycans, that have not been reported to affect αSyn uptake^26^. Taken together, our findings indicate that BAP1 activation reduces HSPG, but not CSPG, levels in cells, which may explain BAP1’s role in regulating αSyn fibril internalization at the plasma membrane.

**Figure 4:**
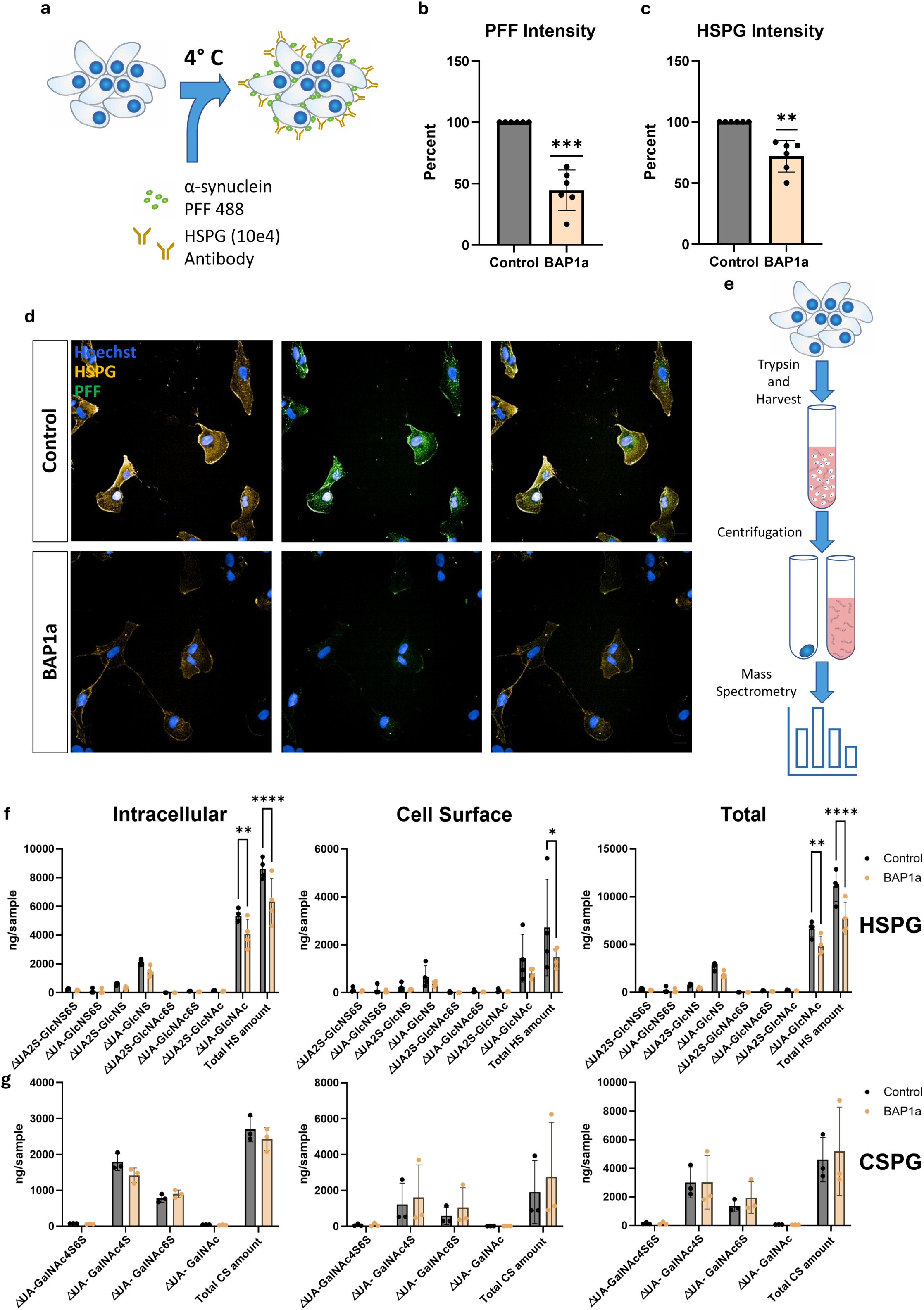
BAP1 activation reduces cell surface HSPG levels and αSyn fibril binding. A) Schematic representation of experiment to investigate cell surface HSPG levels and PFF binding to plasma membrane by arresting endocytosis at 4°C. B) Quantification of mean PFF 488 intensity in BAP1a cells compared to control after endocytosis arrest, representing amount of membrane bound PFF. After background subtraction BAP1a was normalized to its control. n=6 biological replicates. C) Quantification of mean HSPG (10e4) intensity in BAP1 g1 cells compared to control after endocytosis arrest, representing amount of cell surface HSPG levels. After background subtraction BAP1a was normalized to its control. n=6 biological replicates. D) Representative images of PFF binding and HSPG. Hoechst (blue), PFF (green), and HSPG (yellow). Scale bar 20 µm. E) Experimental workflow to measure cell surface and intracellular glycosaminoglycans (HSPG and CSPG) using LC-MS. F) Graphs representing amount of each HSPG-related disaccharide in control and BAP1a cells, separated into intracellular, cell surface (exposed on the extracellular face of the plasma membrane), and total amount of disaccharides in each sample. n=4 biological replicates. G) Graphs representing amount of each CSPG-related disaccharide in control and BAP1a cells, separated into intracellular, cell surface (exposed on the extracellular face of the plasma membrane), and total amount of disaccharides in each sample. n=3 biological replicates. Statistical Analysis: b-c) one sample t-test. f-g) 2-way ANOVA with Šídák’s multiple comparisons test. Graphs are presented as mean ± SD. *p-value<0.05, **p-value<0.01, *** p-value<0.001, ****p-value<0.001

### BAP1 regulates the plasma membrane glycoproteome

As the canonical function of BAP1 involves epigenetic regulation^45^, we asked whether it regulates the cell surface glycoproteome transcriptionally. Using bulk RNA sequencing (RNAseq) we found a large number of differentially expressed genes (DEGs) upon increased BAP1 expression (fig 5a, supplemental table 2). To categorize the DEGs, we performed pathway analysis (DEGs with FDR<0.05 and logFC>|0.75|); strikingly, the top identified terms were associated with the plasma membrane, including extracellular matrix and adhesion (fig 5b). Several genes related to HSPG biosynthesis were found to be differentially regulated, including *EXT1*, *EXTL3*, and *NDST1*, although below our cutoff for transcriptome-wide significance (fig 5c). Moreover, we noticed a group of genes encoding the mucins *MUC3A*, *MUC12*, and *MUC17* that were massively upregulated in BAP1a (fig 5d). The mucin family of proteins are heavily glycosylated through O-linked glycosylation and are the canonical protein carriers for this post translational modifications^46^. Although many are secreted, some mucins including MUC3A, MUC12, and MUC17 are membrane bound and play a role in ligand binding and signalling^46–48^. To validate these findings, we utilized membrane-impermeable NHS-biotin to label the extracellularly exposed cell surface proteome, followed by pulldown with streptavidin beads and mass spectrometry (fig 5e). The isolated fractions showed several cell surface proteins that were differentially abundant in BAP1a compared to control cells (fig 5f, supplemental table 3). Importantly, consistent with our RNAseq data (fig 5d), MUC12 was the most increased cell surface protein (log fold change >4) in BAP1a cells.

**Figure 5:**
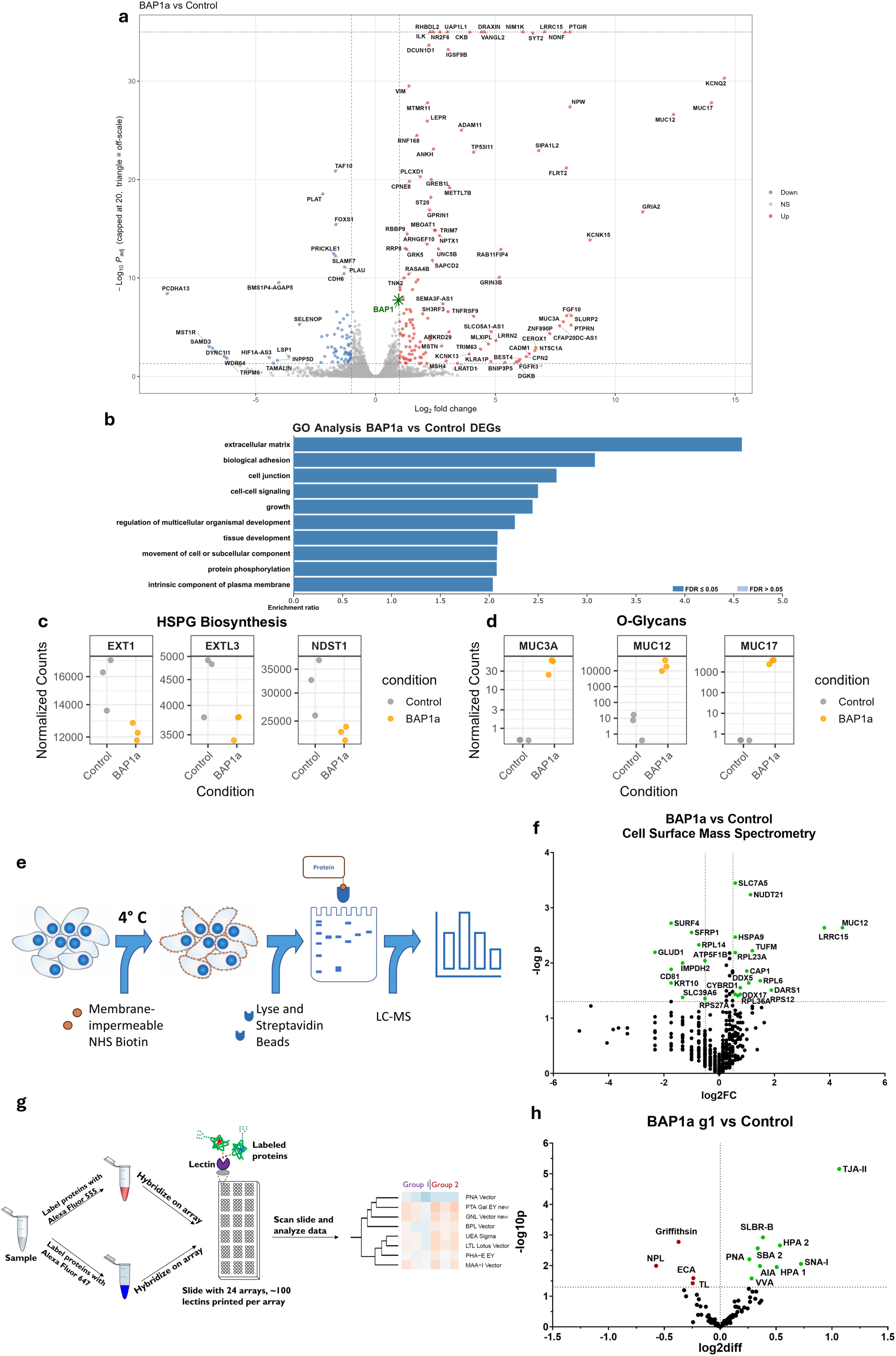
Multi-omic analysis of BAP1a reveals differential regulation of the plasma membrane glycoproteome. a) Bulk RNA sequencing from BAP1a activation cells and CRISPRa controls. Analysis through DEseq2 to display DEGs. n=3 biological replicates. b) Over representation analysis from DEGs within threshold of adjusted p<0.05 and logFC>|0.75| using WebGestalt^40, 41^. c) Normalized counts for HSPG related genes *EXT1*, *EXTL3*, and *NDST1* in BAP1a and CRISPRa controls. d) Normalized counts for O-linked glycoproteins *MUC3A*, *MUC12*, and *MUC17* in BAP1a and CRISPRa controls. e) Experimental workflow for biotin cell surface protein pulldown and mass spectrometry to elucidate changes in the membrane proteome. f) Volcano plot of differentially represented protein form biotin pulldown. Analysis through scaffold, n=3 biological replicates. g) Lectin microarray workflow schematic, used to quantify specific glycan structures on cell membranes, comparing BAP1a and CRISPRa controls. h) Volcano plot of glycan structures of BAP1a compared to CRISPRa controls. For specificity and binding, refer to supplemental figure 9.

As we identified major glycoproteome-related changes in our RNAseq and cell surface proteomics experiments, we examined whether these changes were associated with specific cell surface glycosylation changes that could mediate BAP1’s effect on PFF uptake. To do so, we isolated the plasma membrane from both BAP1a and control cells and analysed them via lectin microarray (fig 5f, supplemental table 4). The lectin microarray quantitatively measures the abundance of lectin-binding moieties, which are the carbohydrate modifications of N-linked and O-linked glycoproteins^49^. Compared to control, BAP1a cells had ∼50% lower binding to mannose lectins (Griffithisin, NPL, and TL), indicating lower N-linked mannose glycan content on the plasma membrane (fig 5g, supplemental table 9). On the other hand, BAP1a had ∼50% increased binding to O-glycan lectins (core 1/3, T/Tn antigen; HPA 1, HPA 2, PNA, and AIA), and their associated α2,3/α2,6-sialylated (SLBR-B and SNA-I) and α2 fucosylated (TJA-II) glycan lectins. Next, to examine whether BAP1 activation led to changes in the subcellular distribution of glycosylation, we labeled cells with fluorescent wheat germ agglutinin (WGA), a pan-glycosylation marker. As glycosylation is most abundant in the Golgi^50^, we compared the co-localization of WGA with the Golgi marker golgin-97 between BAP1a and control cells (fig 6a-b). We noticed not only an increase in the overall levels of WGA staining but a subcellular redistribution of glycosylation with stark reduction in colocalization of WGA and golgin-97 and the appearance of large ridge-like structures in BAP1a cells (fig 6c). Together, these results suggest that BAP1 activation leads to wide ranging transcriptional changes associated with a reorganization of the glycoproteome, involving an increase in cell surface O-glycosylation along with a corresponding increase in expression of mucins, a family of prototypic O-linked glycoproteins. Conversely, we observe a reduction in cell surface high mannose N-glycosylation and HSPG levels as well as a redistribution of glycosylation away from the Golgi.

**Figure 6:**
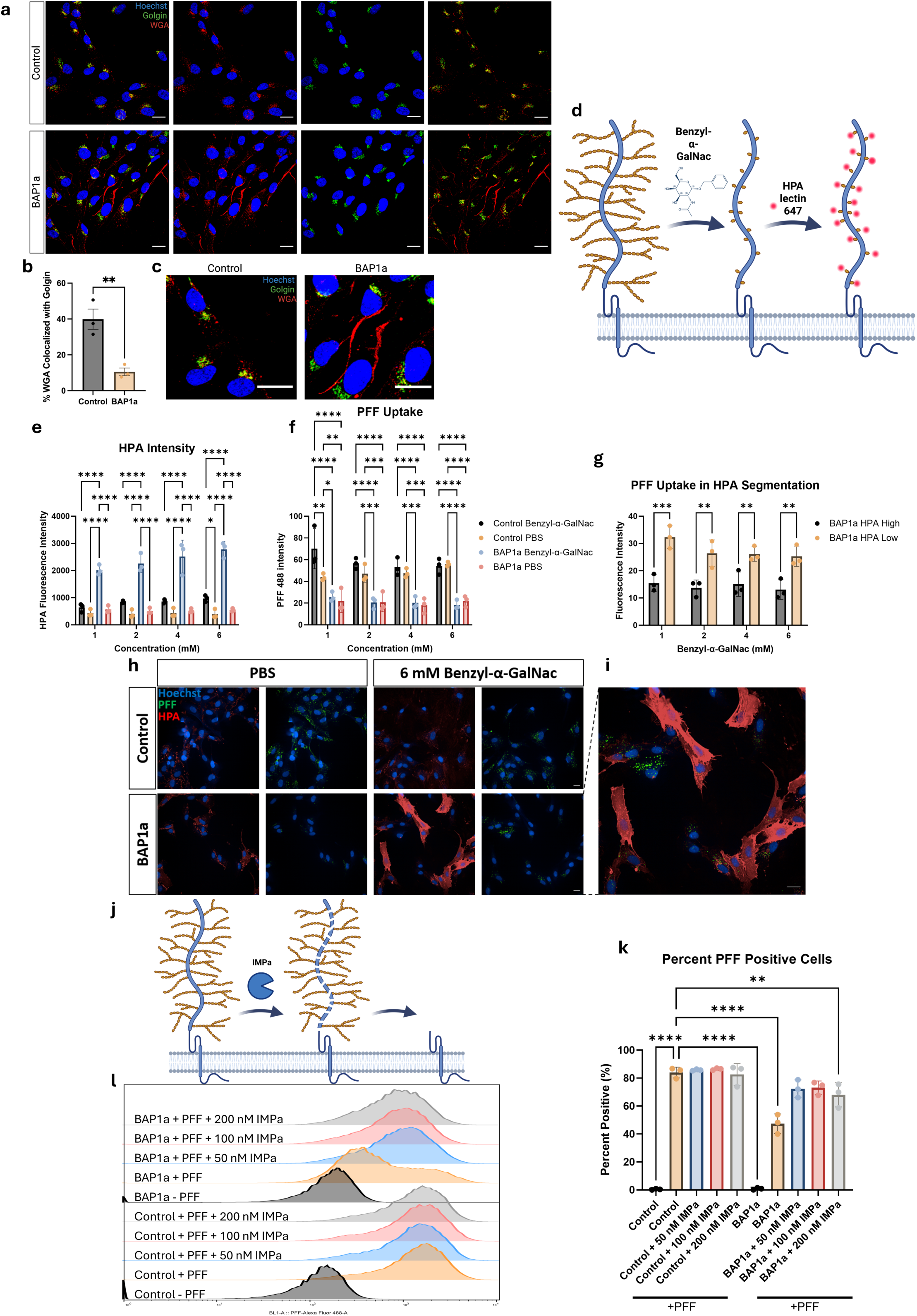
BAP1 regulates αSyn fibril uptake through increased O-linked glycosylation. a) Confocal microscopy of glycans and golgi in CRISPRa control (top) and BAP1a (bottom) cells. Hoechst (blue), golgin-97 (green), Wheat germ agglutinin (WGA; red). Scale bar 20 µm. b) Quantification of percent overlapping signal between WGA and golgin-97. n=3 biological replicates. c) Enlarged view of BAP1a cells compared to CRISPRa control from (a) showcasing distinct ‘ridges’ in BAP1a cells. Scale bar 20 µm. d) Schematic representation of Benzyl-α-GalNac-mediated chain termination of O-glycans and HPA binding to Tn antigen. Created in BioRender. Fon, E. (2026) https://BioRender.com/6x8vl0g. e) Quantification of HPA labelling intensity in control and BAP1a cells treated with benzyl-α-GalNac and vehicle. n=3 biological replicates. f) Quantification of PFF 488 fluorescence intensity in CRISPRa control and BAP1a cells treated with benzyl-α-GalNac and vehicle. n=3 biological replicates. g) Quantification of PFF 488 fluorescence intensity in BAP1a cells treated with benzyl-α-GalNac which display either high or low HPA signal. n=3 biological replicates. h) Representative images of D-F, Scale bar 20 µm. i) Enlarged view of BAP1a cells showcasing low PFF signal in high HPA cells and vice versa. Scale bar 20 µm. j) Schematic representation of IMPa-mediated cleavage of O-glycoproteins before treatment with αSyn fibrils. Created in BioRender. Fon, E. (2026) https://BioRender.com/6x8vl0g. k) Quantification of percentage of PFF-positive cells in CRISPRa control and BAP1a cells pretreated with IMPa. n=3 biological replicates. l) Representative flow cytometry histogram of PFF signal in CRISPRa control and BAP1a cells pretreated with IMPa. n=3 biological replicates. Statistical Analysis: b) two tailed t-test. d-e) 2 way ANOVA with Tukey’s multiple comparisons test. f) 2 way ANOVA with Šídák’s multiple comparisons test.j) one way ANOVA with Dunnett’s multiple comparison test. Graphs are presented as mean ± SD. *p-value<0.05, **p-value<0.01, *** p-value<0.001, ****p-value<0.001

### BAP1 regulates αSyn fibril uptake through increased O-linked glycosylation

We next investigated whether the increase in O-glycosylation *per se* played a role in the reduced internalization of PFFs observed in BAP1a cells. First, we treated cells with Benzyl-α-GalNac, which inhibits the elongation of O-glycosylation chains by blocking chain extension after the first GalNac residue (i.e. exposing the Tn antigen)(fig 6d) ^51, 52^. Treatment with Benzyl-α-GalNac led to a drastic increase in cell surface staining with HPA, a lectin that selectively binds Tn antigen^53^ in BAP1a compared to control cells (fig 6e-f, h). This indicates that BAP1a cells have drastically more cell surface Tn antigen (truncated O-linked glycosylated adducts containing only the first GalNac residue). However, reduction of the O-glycosylation chain-length was not sufficient to reverse the reduced PFF uptake into BAP1a cells (fig 6e-f, h). Exploiting cell-to-cell heterogeneity within the cultures (likely due to differing degrees of BAP1 activation), we found that individual cells with high HPA staining displayed markedly less PFF uptake compared to those with low HPA, indicating that higher levels of cell surface O-linked glycosylation, regardless of chain length, is associated with lower PFF uptake (fig 6g-i).

Since reducing O-linked glycosylation chain-length to Tn antigen did not rescue BAP1’s effect, we explored whether cleavage of cell surface O-glycoproteins could reverse the reduction of PFF uptake into BAP1a cells (fig 6j). We treated cells with IMPa, a protease that cleaves O-glycoproteins specifically at serine/threonine residues^54^, followed by measurement of PFF uptake by flow cytometry. We detected no effect of IMPa on PFF uptake in control cells (fig 6k-l). However, in BAP1a cells, IMPa treatment reversed the reduction of PFF uptake back up to levels observed in control cells (fig 6k-l). Thus, the results support a critical role for BAP1-induced cell surface O-glycosylation, regardless of chain-length, in reducing PFF uptake into cells. Taken together with its effects in reducing HSPGs, our findings support a critical role for BAP1 in regulating αSyn fibril internalization by remodelling the cell surface glycoproteome.

### BAP1 modifies αSyn fibril internalization in dopamine neurons but not microglia

PD pathology is characterized by the loss of DNs in the SN^1^. We thus sought to determine whether BAP1 also controls PFF uptake in DNs. A human iPSC line from a healthy individual harbouring inducible NGN2 and the CRISPRa machinery^55^ was transduced with either BAP1a or control non-targeting sgRNAs, and differentiated into DNs (iDAs), as previously described^56–58^, followed by qPCR to confirm *BAP1* overexpression (Supplemental fig 3a-b). Similar to our results in RPE-1 cells, BAP1a iDAs exhibited a reduction in PFF uptake compared to the control iDAs (fig 7a,b). A hallmark of PD is the aggregation of αSyn, which can be monitored by the appearance of phosphorylated αSyn (pSyn) in brain cells^59, 60^. This can be modeled experimentally in DNs by brief incubation (24 hour) with PFFs followed by long-term culture (≥3 weeks) to induce the phosphorylation of endogenous αSyn at serine 129^16, 17, 59, 60^ . Compared to control iDAs, we observed a reduction in pSyn puncta in BAP1a iDAs (fig 7c,d). Next, to determine whether the effect of BAP1 was specific to DNs or also regulated αSyn fibril internalization in other PD-relevant cell types, a human iPSC line from a healthy individual harbouring six doxycycline-inducible microglial transcription factors, as well as the CRISPRa machinery (6TF-iMG)^61^ was transduced with either BAP1a or control non-targeting sgRNAs, and differentiated into microglia as described^61^, followed by qPCR to confirm *BAP1* overexpression (Supplemental fig 3c-d). In contrast to iDAs, BAP1a 6TF-iMGs, staining positive for the classical microglia marker Iba1, showed no significant differences in PFF uptake compared to control 6TF-iMGs (fig 7e-h). These results suggest that BAP1 selectively regulates PFF internalization in DNs but not microglia.

**Figure 7:**
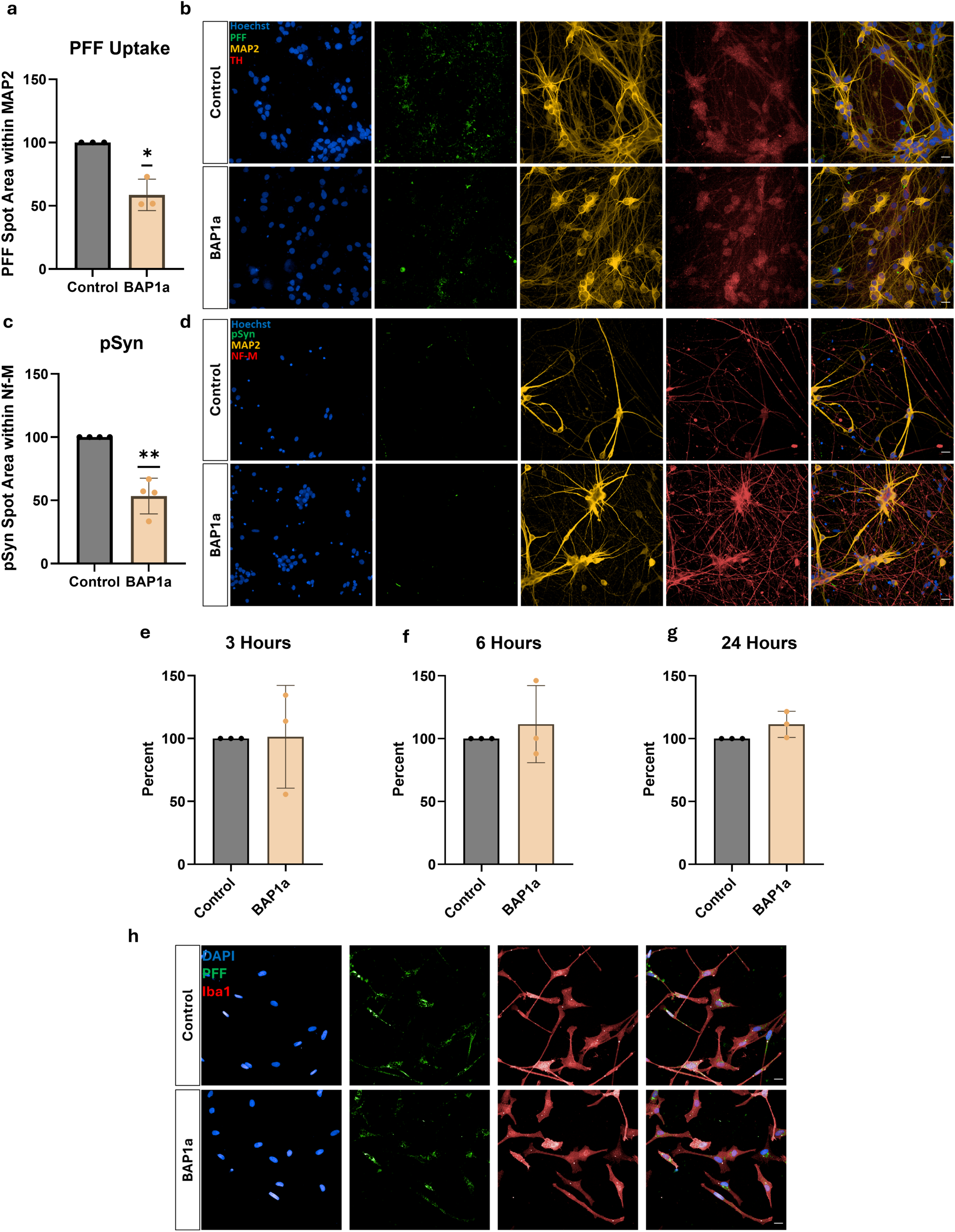
BAP1 regulates αSyn fibril internalization and seeding in dopamine neruons but not microglia. a) PFF uptake assay in iDAs. Quantification of PFF 488 spots within defined MAP2 comparing BAP1a to CRISPRa control after 24 hours of PFF treatment. BAP1a normalized to control within each biological replicate. n=3 biological replicates. b) Representative images of PFF uptake assay in iDAs. Hoechst (blue), PFF (green), MAP2 (yellow), TH (red). Scale bar 20 µm. c) αSyn seeding assay in iDAs. Quantification of phosphorylated αSyn (pSyn) within neurofilament-medium stain comparing BAP1a to sgRNA controls after 24-hour PFF treatment and 3-week seeding. n=4 biological replicates. d) Representative images of αSyn seeding assay in iDAs. Hoechst (blue), pSyn (green), MAP2 (yellow), NF-M (red). Scale bar 20 µm. e) PFF uptake assay in 6TF-iMG. Quantification of PFF 488 intensity in Iba1 cells, comparing BAP1a to sgRNA control after 3 hours of PFF treatment. BAP1a normalized to control within each biological replicate. n=3 biological replicates. f) PFF uptake assay in 6TF-iMG. Quantification of PFF 488 intensity in Iba1 cells, comparing BAP1a to sgRNA control after 6 hours of PFF treatment. BAP1a normalized to control within each biological replicate. n=3 biological replicates. g) PFF uptake assay in 6TF-iMG. Quantification of PFF 488 intensity in Iba1 cells, comparing BAP1a to sgRNA control after 24 hours of PFF treatment. BAP1a normalized to control within each biological replicate. n=3 biological replicates. h) Representative images of PFF uptake assay in 6TF-iMG after 3 hours of PFF treatment. Hoechst (blue), PFF (green), Iba1 (red). Scale bar 20 µm. Statistical Analysis: one sample t test. Graphs are presented as mean ± SD. *p-value<0.05, **p-value<0.01, *** p-value<0.001, ****p-value<0.001

### BAP1 and ASXL3, but not ASXL1 and ASXL2, regulate αSyn fibril internalization and dopamine neuron vulnerability in a midbrain organoid model of Parkinson’s Disease

BAP1’s function has largely been attributed to its role in the PR-DUB^30, 31, 45, 62, 63^. This complex is comprised of several proteins, including OGT and HCF1; however, some of the best documented binding partners of BAP1 within the PR-DUB complex are Additional Sex Combs Like (ASXL) proteins. ASXL1, ASXL2, and ASXL3 are mutually exclusive within the PR-DUB complex, and the presence of each dictate the genomic targets, and thus, where BAP1 acts. The expression of the ASXLs is different across cell types; ASXL3 in particular has been reported to be important to the brain, and it’s expression is higher in neurons compared to other cell types in the brain (BRAIN-seq)^34^. We hypothesized that this differential cell expression and activity may underly the cell-specific effect that we observed with regards to αSyn fibril internalization between DNs and microglia, especially as a result of a shifted stoichiometric balance between BAP1 and the ASXLs. Thus, we analyzed data from generated iPSC-derived three-dimensional assembloids consisting of human midbrain organoids (hMOs) containing integrated microglia (intMG) to confirm the distribution of the respective *ASXL*s and *BAP1* across various PD-relevant brain cell types^64–66^. Using single cell RNA sequencing (scRNAseq)(fig 8*a*), we found that the expression levels of most genes of interest (*BAP1*, *ASXL1*, and *ASXL2*) did not meet the threshold for significance (FDR<0.05 and logFC>|1|) between DN and microglial populations (fig 8b-d, supplemental table 5). In contrast, *ASXL3* expression was much lower in the microglial population compared to most neuronal subtypes, and especially the DN population (FDR=4.9x10^-65^, logFC= -3.3) (fig 8*e*). This pattern of *ASXL* distribution across the different cell types in assembloids suggests that presence of ASXL3 within the PR-DUB complex could mediate BAP1’s effect on remodelling the cell surface glycoproteome and its downstream effect on αSyn fibril internalization.

**Figure 8:**
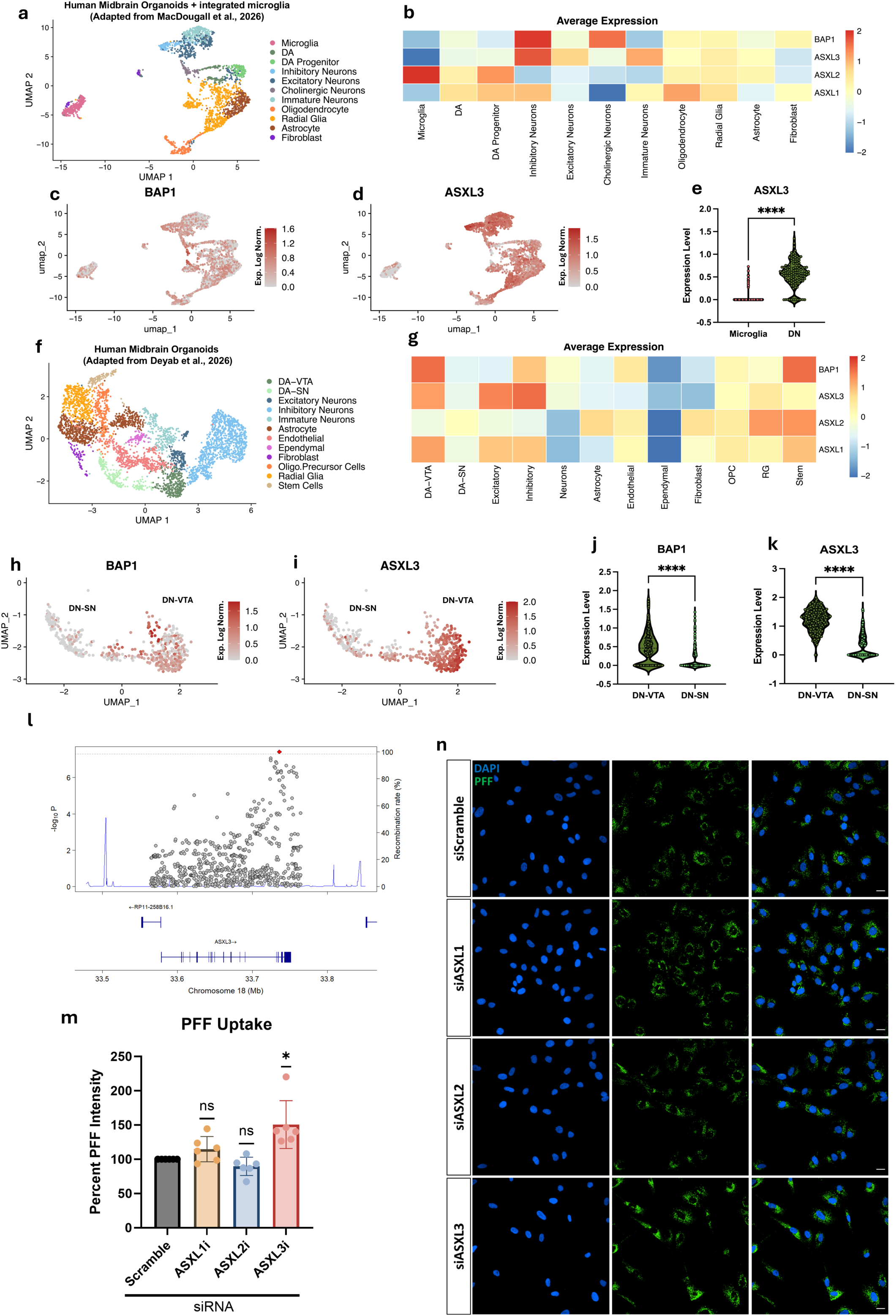
*BAP1* and *ASXL3* regulate αSyn fibril internalization and dopamine neuron vulnerability in a midbrain organoid and assembloid models of PD. a) scRNA UMAP showing cell types in assembloids (hMO + intMG), adapted from MacDougall et. al., 2026^64^. b) Heatmap representing normalized average expression of *BAP1*, *ASXL1*, *ASXL2*, and *ASXL3* in cell types found in hMO + intMG. c) scRNA UMAP showing distribution of *BAP1* expression across cell types in hMO + intMG. d) scRNA UMAP showing distribution of *ASXL3* expression across cell types in hMO + intMG. e) Violin plot comparing *ASXL3* expression in dopaminergic neurons and microglia in hMO + intMG. f) scRNA UMAP showing cell types in SNCA triplication and isogenic control hMOs adapted from Deyab et. al., 2026^69^. g) Heatmap representing normalized average expression of *BAP1*, *ASXL1*, *ASXL2*, and *ASXL3* in cell types found in SNCA triplication and isogenic control hMOs. h) scRNA UMAP showing distribution of *BAP1* expression across dopaminergic subtypes in SNCA triplication and isogenic control hMOs. i) scRNA UMAP showing distribution of *ASXL3* expression across dopaminergic subtypes in SNCA triplication and isogenic control hMOs. j) Violin plot comparing *BAP1* expression between DN-VTA (protected) and DN-SN (vulnerable) in SNCA triplication and isogenic control hMOs. k) Violin plot comparing *ASXL3* expression between DN-VTA (protected) and DN-SN (vulnerable) in SNCA triplication and isogenic control hMOs. l) LocusZoom plot displays PD GWAS association across the ASLX3 region^10^. m) PFF uptake assay on ASXL1-3 siRNA-mediated knockdown. Quantification of PFF 488 intensity in cells, normalized to control in each experiment. n) Representative images of ASXL1-3 knockdown exhibiting increased PFF signal in *ASXL3* knockdown compared to the siScramble control. Hoechst (blue) and PFF (green). Scale bar 20 µm. Statistical Analysis: two tailed t test. Graphs are presented as violin plots, with each point representing a single cell. *p-value<0.05, **p-value<0.01, *** p-value<0.001, ****p-value<0.001

Next, we explored whether the cell-type distribution of *BAP1* and the respective *ASXLs* within hMOs was associated with regional vulnerability in PD. Using scRNAseq in 5-month old hMOs derived from a PD patient with a triplication of the *SNCA* gene^67, 68^, we reported a marked reduction in a subpopulation of DNs, compared to isogenic CRISPR-corrected control hMOs from the same patient^69^. The subpopulation that is lost in the *SNCA* triplication closely resembles the DNs of the SN (DN-SN), whereas the DNs that survive resemble the DNs of the ventral tegmental area (DN-VTA), which is less vulnerable to degeneration in PD^69^ (fig 8f; light-and dark-green, respectively). Upon examining of *BAP1*, *ASXL1*, *ASXL2*, and *ASXL3* expression in these hMOs, we identified a marked reduction in the expression of *BAP1* and *ASXL3* in the DN-SN compared to DN-VTA that met the DEG cutoff (FDR<0.05 and logFC>|1.5|) (fig 8h-k, supplemental table 6). These data indicate that elevated expression of both *BAP1* and *ASXL3* is associated with more resilient subpopulations of DNs, whereas lower expression is associated with DN subpopulations that are more vulnerable to degeneration in PD, similarly to patterns shown in fig 2a. Moreover, its nomination in a recent PD GWAS (fig 8l) and the identification of rare loss-of-function variants in PD patients^12, 70^, further support a role for *ASXL3* in PD genetics and pathogenesis.

Finally, we empirically tested whether a reduction in the ASXL3 could phenocopy the effect of *BAP1* reduction (fig 3i-k) and lead to an increase in αSyn fibril internalization. Using siRNA to knockdown *ASXL1, ASXL2,* or *ASXL3* in RPE1 cells (supplemental fig 3e-g), followed by incubation with PFFs, we found that reduction of ASXL3, but not ASXL1 or ASXL2, leads to increased PFF uptake (fig 8m-n). Taken together, these results indicate that BAP1 and ASXL3, both PD risk genes, regulate αSyn fibril internalization epigenetically via their binding in the PR-DUB complex. Moreover, this effect is cell-specific likely due to the varying expression of *ASXL3* in neurons compared to microglia. In this scenario, the low levels of *ASXL3* expression in microglia could explain why increasing *BAP1* expression alone via CRISPRa (but not *ASXL3*) is insufficient to reduce PFF uptake.

## Discussion

Membrane glycosylation, namely in the form of HSPGs, has been shown previously to play a role in the internalization of αSyn aggregates, providing a molecular mechanism to explain the cell-to-cell spread of pathology in PD^26–29^. However, the association between cell surface glycosylation pathways and PD genetics has been less clear^10–12^. In this study, we report that *BAP1* and *ASXL3*, two PD risk genes that function in the PR-DUB complex, function as regulators of αSyn fibril internalization via cell surface glycosylation. Our findings showing BAP1-induced transcriptional changes in known cell membrane glycosylation carrier proteins is consistent with the known function of the PR-DUB in the epigenetic regulation of chromatin accessibility leading to transcriptional changes in multiple genes^32, 62, 71^. Indeed, mutations in *BAP1* have been reported to change the membrane glycoproteome via transcriptional regulation in both breast and clear cell renal carcinoma, and this effect is largely attributed to its deubiquitinase activity^71, 72^. Here we show that this effect is also relevant for PD, as the cell-surface glycosylation changes induced by BAP1 regulate αSyn fibril internalization. Moreover, whereas both *BAP1* and *ASXL3* have been mapped to PD GWAS risk loci^11, 12^ and rare loss-of-function mutations in *ASXL3* have been associated with PD^70^, here we show, using colocalization and Mendelian randomization, that reduced *BAP1* expression is associated with increased PD risk.

Regulation of the membrane glycoproteome by BAP1 provides insights into known and novel mechanisms of αSyn fibril internalization. We find that higher BAP1 expression decreases cell surface HSPG content, which in turn reduces binding of PFFs. This agrees with previous studies that explore the role of HSPG in αSyn binding^26–29^. Beyond αSyn fibrils, HSPGs have also been implicated in viral entry into cells, mediating the binding of viral particles through HS’ chain negative charge^73, 74^. Indeed, several viruses have been shown to bind and enter cells through HSPGs, including SARS-CoV-2, which binds HSPGs through its Spike proteins^75^. The parallels between HSPG-mediated cellular internalization of viruses and αSyn fibrils have been proposed previously^75, 76^. Moreover, other forms of glycosylation have also been implicated in virus internalization, including O-glycosylation^77, 78^. Given these similarities, it would be interesting to determine whether BAP1-mediated reprogramming of the cell-surface glycoproteome can also regulate viral entry into cells, as we have shown for αSyn fibrils.

Indeed, our multi-omics approach showed BAP1-mediated reduction in HSPG biosynthetic enzymes, and an increase in cell surface O-linked glycosylation carrier proteins such as mucins. O-linked glycoproteins have yet to be described to play a role in αSyn internalization, despite being abundant in brain^79–81^. Forming protective barriers and playing important roles in cell signalling and cargo binding, dysregulation of O-glycans has been suggested to be involved in brain aging^81, 82^. We find that, in conjunction with HSPG, BAP1-mediated increased O-glycosylation is responsible for BAP1’s control of αSyn internalization as cleavage of O-glycans reverses BAP1’s effect on αSyn uptake. This provides a novel two-pronged mechanism involving the regulation of cell surface HSPGs and O-linked glycoproteins in opposing directions via the transcriptional effects of BAP1. Interestingly, Shi and colleagues (2025) show that aging leads to reduced O-glycans in the blood-brain-barrier in mice^82^, further suggesting a role for O-glycans in the age-related increase in the spread of pathogenic proteins such as αSyn spread into the CNS from the rest of the body. Indeed, gut-to-brain transmission of αSyn has been shown in several models of PD^83, 84^, warranting further studies to determine whether O-glycans and their regulations are involved.

In PD, the main cell loss occurs in DNs within the SN of the midbrain^1^. The loss of these neurons is widely thought to occur due to αSyn accumulation and aggregation, which has been shown in several models^2, 16, 17^. Pathologically, different brain regions have differential αSyn expression levels and differential vulnerabilities to PD pathology^42, 43^. Moreover, the involvement of microglia, the immune residents of the brain, has been shown to be important for the spread of αSyn throughout the brain^85–88^. We show that activation of BAP1 in iPSC-derived dopamine neurons, but not microglia, reduced αSyn PFF uptake and aggregation. The role of BAP1 as a histone deubiqitinase (DUB) has been reported to be cell-type specific, based on the mutually exclusive PR-DUB complex it forms with the scaffolding protein ASXL1, ASXL2 or ASXL3^32, 33, 89–91^. The expression of these differs between cell types. In particular, *ASXL3* is highly expressed in the brain (particularly in neurons)^34^ and its mutation causes the neurodevelopmental disorder, Bainbridge-Ropers syndrome^92^. Using a 3D human organoid model, we show that *ASXL3* is expressed highly in neurons (especially DNs) but not in microglia, consistent with the model whereby the lack of ASXL3 in microglia explains the lack of effect of *BAP1* on αSyn fibril internalization in these cells. Indeed, knockdown of ASXL3, but not ASXL1 or ASLX2, increased αSyn PFF uptake, phenocopying the effects of BAP1 knockdown and suggesting they are both required within the PR-DUB complex to regulate αSyn fibril internalization.

Degenerating neurons in SN are particularly vulnerable to death in PD, whereas their VTA counterparts are generally believed to be more resilient^35, 43, 93, 94^. The notion of selective vulnerability is central to PD; importantly, several studies identify DN subtypes within the SN and VTA that are differentially vulnerable in PD^35, 36^. Using an *SNCA* gene triplication as a model for region-specific neurodegeneration, we mapped the expression patterns of vulnerable and resistant neurons within 3D organoids that correspond to SN-like and VTA-like DNs. Our previous studies show that this model can be used to study selective neuronal loss associated with αSyn pathology^69^. Here, we show that the expression of *BAP1* and *ASXL3* was significantly increased in the VTA-like cells compared to SN-like DNs, suggesting that increased *BAP1*-ASXL3 axis is associated with protection in synucleinopathy. The regional expression pattern of *BAP1* has been found to be inversely associated with vulnerable brain regions in PD according to Braak’s staging paradigm, with its expression being the highest in the cortex, and its expression is the lowest in the SN^42^, consistent with the mechanisms we report here of reduced αSyn fibril internalization with higher *BAP1* expression. Taken together, the findings support both a genetic and functional role for BAP1/ASXL3-containing PR-DUB complexes in determining cell-type vulnerability in PD by reprogramming the surface glycoproteome to regulate αSyn fibril uptake into cells.

## Methods

### Cell culture

RPE1 and HEK293T (for lentivirus generation) cells were cultured using complete Dulbecco’s Modified Eagle Medium (DMEM, supplemental table 8). Passaging the cells included a brief wash with sterile phosphate buffered saline (PBS), followed by detachment with trypsin/EDTA (Wisent) at 37 °C until optimal dissociation followed by resuspension in complete medium. Cells were then plated to the desired density after counting. Cells were always maintained at 37 °C and 5% CO_2_ and routinely tested for mycoplasma using a PCR detection kit (ABM).

### RPE1 Cell Line Generation

sgRNA sequences were synthesized as oligonucleotides (IDT), annealed, and cloned into a BstXI/BlpI digested pCRISPRia-v2 plasmid (addgene #84832), as detailed^37^. Lentivirus was then generated and used to infect cells at a low MOI with polybrene (8 μg/mL). RPE1 cells were then selected and/or sorted into single cells by FACS based on BFP fluorescence to generate polyclonal cell lines.

### CRISPRa Screen

RPE1 CRISPRa cells were infected with hCRISPRa-v_2_^38^ lentiviral library at a MOI<0.3 in suspension with 8 µg/mL polybrene, and seeded at 5x10^6^ cells per 15 cm dish (library coverage at infection ∼500x). Media was replaced 24 hours after infection, allowed to recover for another day, and selected with puromycin (15 μg/mL) for 72 hours. After selection, cells were expanded for 48 hours, then replated at 5x10^6^ cells per 15 cm dish. Cells were then treated with 15 nM PFF-488 for 24 hours before trypsinization and harvesting. Cells were then fixed in suspension with 4% PFA in PBS for 15 minutes, before sorting on BD Biosciences FACSAria Fusion; only BFP positive cells were selected and separated into the top and bottom 15% of cell distribution in terms of PFF fluorescence (library coverage ∼115x-130x, determined by sorted cell numbers in each bin). Genomic DNA was extracted as previously described. A stepwise PCR was performed to amplify the sgRNA region (primers shown in supplemental table 7); the outer PCR was performed with 2X KAPA HiFi HotStart ReadyMixPCR Kit and run on a 2% agarose gel. 600 bp band was then isolated with QIAquick Gel Extraction Kit (Qiagen) and used as the template for the inner PCR. The inner PCR fragment (∼200 bp) was also excised from an agarose gel and sequenced on Illumina NextSeq 500. The data was then analyzed using MAGeCK^39^.

### Genetic Analysis

#### Brain expression, eQTL, and genetic analyses

Bulk tissue expression of *BAP1* across human brain regions was examined using GTEx v10. *BAP1* expression across brain tissues was visualized to compare regional expression patterns, including cortex, cerebellum, substantia nigra, and basal ganglia regions.

Brain eQTL associations at the *BAP1* locus were examined using BrainMeta v2 cortex cis-eQTL summary statistics (N = 2,443)^95^. Regional PD GWAS and *BAP1* eQTL association signals were compared across the *BAP1* locus. Linkage disequilibrium between rs123598 and rs34332947 was assessed using the 1000 Genomes Project Phase 3 European reference panel. Genotype-associated *BAP1* expression in caudate, cortex, hippocampus, and nucleus accumbens was examined for rs123598 and rs34332947 using GTEx v10.

#### Summary-data-based Mendelian randomization

Summary-data-based Mendelian randomization (SMR)^96^ was performed using BrainMeta v2 cortex cis-eQTL summary statistics for *BAP1* (N = 2,443)^95^ as the exposure and PD GWAS summary statistics comprising 63,555 cases, 17,700 proxy cases, and 1,746,386 controls^10^ as the outcome. Significant cis-eQTLs (P ≤ 5 × 10⁻⁸) within ±500 kb of *BAP1* were considered for analysis. The HEIDI test was used to assess heterogeneity in the association signal and to distinguish a shared pleiotropic signal from associations arising through linkage.

#### Colocalization analysis

Colocalization analysis was performed between GP2 PD GWAS summary statistics^10^ and BrainMeta eQTL summary statistics^95^ at the *BAP1* locus (±100 kb) using coloc R package with the SuSiE framework (coloc.susie)^97^, which allows for multiple causal signals within a locus. SuSiE fine-mapping was performed using summary statistics and an LD reference matrix. Posterior probabilities were calculated for the five standard colocalization hypotheses, including PP.H4, which represents the probability that the GWAS and eQTL signals share a causal variant. A PP.H4 > 0.8 was considered evidence of strong colocalization.

#### Genetic Data availability

BrainMeta v2 cis-eQTL summary statistics are publicly available through the SMR data resource at https://yanglab.westlake.edu.cn/software/smr/#DataResource. Full PD GWAS summary statistics from GP2 are publicly available through the AMP-PD/NDKP data portal at https://ndkp.hugeamp.org/research.html?pageid=a2f_downloads_280^10^.

#### Western Blot

Cells are pelleted and lysed in RIPA buffer for 30 minutes, before sonicating for 10 seconds at 40%. Samples are then centrifugated to remove debris, and the protein concentration is measured by Pierce BCA assay (ThermoFisher). After diluting samples as needed with 6X Laemmli buffer, samples are boiled at 95 °C for 5 minutes, centrifugated, and loaded onto SDS-PAGE gel. Samples were then transferred onto nitrocellulose membranes by semi-dry turbo transfer (as recommended by pre-set settings, Bio-Rad). After blocking in 5% skim milk in PBS-T, membranes were incubated overnight with antibody of interest in blocking solution. Then, membranes were washed thrice with PBS-T and incubated with respective secondary antibody in blocking solution for 1 hour. Membranes were washed thrice before imaging on ChemiDoc, using either Clarity or ClarityMax ECL substrate (BioRad).

#### Real Time Quantitative PCR (RT-qPCR)

RNA was extracted using Trizol-chloroform method and purified using RNeasy kit (Qiagen). 0.5-2 μg RNA were then used to synthesize using M-MLV reverse transcriptase kit (Invitrogen). The cDNA was diluted before assembling PCR reactions with SsoAdvanced master mix (BioRad) with designed primers and run on QuantStudio 3 (ThermoFisher).

#### Cloning

sgRNAs were synthesized as oligonucleotides with complementary overhangs for BstXI/BlpI restriction digests (IDT). Oligos were then resuspended, annealed, and digested with BstXI/BlpI, before ligating into a BstXI/BlpI digested pCRISPRi/a-v2 plasmid.

#### αSyn Purification and PFF Generation

PFFs were made from recombinant αSyn monomers, as previously described^98, 99^. The labelling of PFF with pHrodo green dye (Thermo Fisher Scientific) was performed following the manufacturer’s instructions.

#### High Throughput PFF Uptake Assays

Cells were seeded in 96 well plates in desired density a minimum of 24 hours prior to treatment or fixation. For PFF internalization assays, cells were treated in complete media with 80 nM Alexa488-PFFs for 3, 6, or 24 hours as indicated before fixation with 4% paraformaldehyde (PFA). Hoechst (1/10000) was used to stain nuclei, and phalloidin-TRITC was used to image cell bodies to define cells. For siRNA experiments, cells were reverse transfected using HiPerfect (Qiagen) 48 hours prior to PFF treatment. Imaging was performed on Perkin Elmer Opera Phenix, and analysis was done using Perkin Elmer Columbus using published pipelines.

#### Flow cytometry PFF Uptake

Cells were seeded in desired density in 12 well plates a day before PFF treatment. After treatment with 15 nM Alexa488-PFFs for 3 hours, cells were washed, trypsinized, and pelleted before fixation with 4% PFA in suspension. After washing with PBS, cells were analyzed for PFF fluorescence using ThermoFisher Attune NxT Flow Cytometer and analyzed using FloJo.

#### PFF Binding and HSPG Staining

RPE1 cells seeded in a 96 well plate were washed with cold serum-free DMEM and placed on ice for minimum 5 minutes. Then cells were incubated with 180 nM PFF-488 and 1/500 anti HSPG antibody (10e4, Amsbio) for 20 minutes on ice. After washing in cold serum free DMEM, cells were fixed with 4% PFA for 15 minutes, rinsed with PBS, and blocked with 5% normal goat serum solution. Finally, the cells were stained with 1/1000 anti-mouse 555 secondary antibody (ThermoFisher), 1/1000 wheat germ agglutinin 647 (ThermoFisher), and 1/5000 Hoechst for 25 minutes, and rinsed before imaging on the Opera Phenix.

#### O-Glycan Chain Reduction Benzyl-alpha-GalNac

Cells were seeded at low density in 96 well plates for 24 hours, before treatment with PBS, 1, 2, 4, 6 mM of Benzyl-alpha-GalNac in complete media for 72 hours. Cells were then treated with 80 nM of PFF for 3 hours and fixed as described above. We then stained the cells with in-house 647-conjugated HPA lectin (5 µg/mL) for 2 hours at room temperature, as well as Hoechst and phalloidin 555. Imaging was done on Opera Phenix and analyzed using Columbus.

#### Bulk RNA Sequencing

Control and BAP1a g1 cells were grown in 10 cm dishes until 80% confluency before extraction as previously described (see RT-qPCR). RNA was quantified on a nanodrop before polyA enrichment and sequencing using NovaSeqPE. Analysis was done using DEseq2 on Rstudio for differentially expressed genes.

#### Cell Surface Proteomics

RPE1 cells were seeded in 10 cm dishes and grown to ∼90% confluence. Each plate was washed with PBS (with Ca/Mg, Wisent), and incubated with 0.25 mg/mL EZ-Link™ Sulfo-NHS-Biotin (ThermoFisher, A39256) solution for 30 minutes at 4°C. After another wash with PBS, unbound biotin was deactivated with 100 mM glycine for 15 minutes. After washing, cells were scraped and pelleted and frozen at -80°C. Cells were then lysed with 10 mM Tris-HCl, 150 mM NaCl, 1% Triton, 0.1% SDS and 1mM protease inhibitor cocktail for 30 minutes. After 5 second sonication, cells were centrifuged to remove debris, and incubated with 70 μL Pierce™ High Capacity Streptavidin Agarose (ThermoFisher, 20357) for 3 hours. Samples were then analyzed by LC-MS at The Proteomics and Molecular Analysis Platform of McGill University Health Centre Research Institute and quantified in Scaffold.

#### Lectin Microarray

BAP1 g1 and CRISPRa control cells were seeded at 5x10^6^ cells per 15 cm dish for 24 hours. Cells were then rinsed with PBS, incubated with 0.5M EDTA (pH = 8) for 15 minutes at room temperature, and scraped. After further scraping with PBS, cells were pelleted and frozen at -80 °C. Cell membranes were then isolated by sonication and ultracentrifugation at 100,000 g for 1 hour before proceeding with protein quantification, labelling, and lectin microarray analysis as described previously^49, 100^. Details of the print and lectin microarray analysis can be found in the MIRAGE table (Supplemental table 9).

#### iPSC-derived dopamine neurons

Briefly, WTC11 harbouring doxycycline-inducible NGN2 and trimethoprim (TMP)-inducible CRISPRa iPSCs obtained from Dr. Martin Kampmann were cultured at 37°C in supplemented E8. For BAP1 activation in iPSCs, the NGN2-CRISPRa iPSCs were transduced at the iPSC stage using BAP1a g1 lentivirus and selected with puromycin. Protocol is largely based on Sheta and colleagues^56^.

To differentiate iPSCs into neuronal progenitor cells (NPCs), iPSCs were dissociated using Accutase and transferred to Matrigel-coated 6-well plates (1 million cells per well) in predifferentiation media for 3 days. For differentiation of early NPCs into dopaminergic neurons, the cells were dissociated using Accutase and plated in either 96-well plates (15,000 cells per well) or 6-well plates (750,000 cells per well) on polyornithine/laminin-coated surfaces in Neuronal Differentiation Media. After 3 days, media was fully replaced by dopaminergic maturation media supplemented with TMP to induce CRISPRa. For seeding assays, neurons were treated 3 days later with either 0.3 µM of PFF or PBS, and cultured for 4 weeks. For PFF uptake assays, cells were cultured for 2 weeks before treatment with 80 nM PFF-488 for 24 hours. Dopaminergic neurons were maintained by exchanging half of the culture volume for fresh final dopaminergic differentiation media every 5 days. For media formulations, see supplemental table 8.

For uptake assays, neurons were stained with Hoechst, MAP2, and TH; for seeding assays neurons were stained with Hoechst, MAP2, and Neurofilament-M (Nf-M). Imaging was performed on Opera Phenix and analyzed in Columbus.

#### iPSC-derived Microglia

iTF-Microglia were differentiated as described by Dräger and colleagues^61^. The CRISPRa iTF-iPSC line with doxycycline-inducible expression of microglial factors PU.1, CEBPβ, IRF5, MAFB, CEBPα and IRF8, were a gift from Dr. Martin Kampmann. Briefly, iTF-iPSC were cultured in StemFlex Basal Medium on Matrigel coated dishes, with medium replacement every 1-2 days, as necessary. CRISPRa iTF-iPSC were passaged using Accutase and cultured with 10 mM Y-27632 ROCK inhibitor for 24 hours after passaging. CRISPRa iTF-iPSC were transduced with lentivirus encoding individual sgRNAs targeting BAP1, selected with 2 µg /ml puromycin for 2–4 days, and allowed to recover in the absence of puromycin for 2-4 days. To generate CRISPRa iTF-Microglia, CRISPRa iTF-iPSCs were dissociated using Accutase, counted and seeded in day 0 onto plates doubled coated with Poly-D-Lysine and Matrigel at a density of 10,000 cells per well in a 96-well plate or 150,000 cells per well in a 6-well plate. On day 2, medium was replaced with day 2 medium. On day 4, medium was replaced with iTF-Microglia media day 4. A full medium change to fresh iTF-Microglia medium was performed every 3-4 days until cells were assayed on day 10. All media formulations are outlined in supplemental table 8.

## Data Availability

All genetic data is openly available from GTEx (v10; https://gtexportal.org/home/), BrainMeta v2 (https://yanglab.westlake.edu.cn/software/smr/#DataResource), and GP2 Parkinson's disease GWAS available through the AMP-PD/NDKP data portal (https://ndkp.hugeamp.org/research.html?pageid=a2f_downloads_280). All other data and materials in the present work are available upon reasonable request to the authors.

## Acknowledgements

We thank Wolfgang Reintsch for maintenance and technical support of the high content imaging facility, and the Early Drug Discovery Unit (EDDU) at McGill University for supporting and providing technical assistance with high content imaging, flow cytometry, and FACS. We also thank the Proteomics and Molecular Analysis Platform at McGill University and Donnelly Sequencing Centre at University of Toronto for their technical assistance.

---

**Supplemental figure 1:**
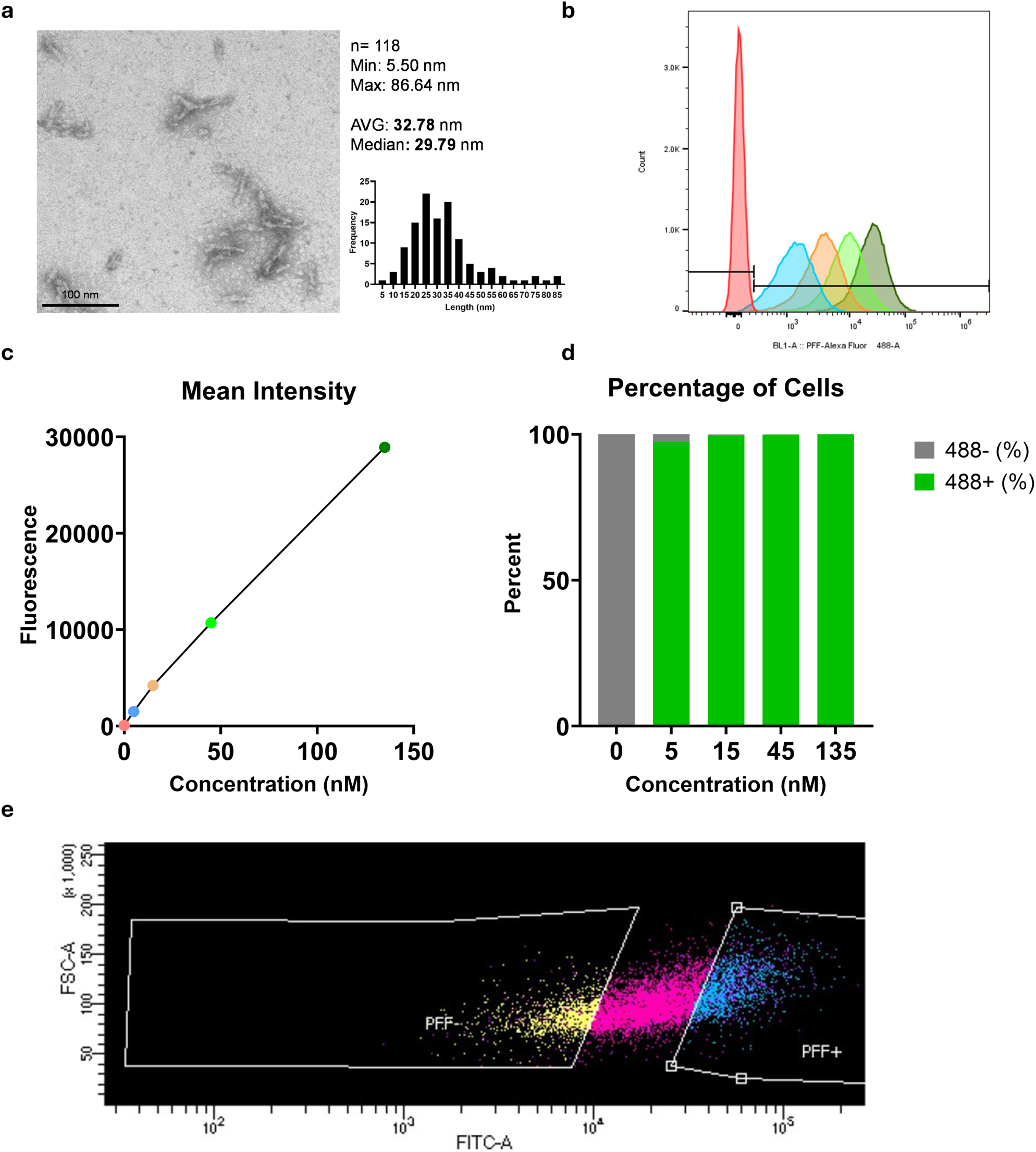
Quality control and internalization of αSyn PFF. a) Electron microscopy of sonicated αSyn PFF that was used for the screen and quantification of size and distribution. b) Flow cytometry histogram showing fluorescence of PFF 488 after 24 hour treatment of increasing concentrations of PFF. 0 (red), 5 (turquoise), 15 (orange), 45 (light green), 135 (dark green) nM PFF. c) Quantification of mean fluorescence intensity of increasing PFF concentration as shown in b. d) Quantification of percentage of PFF-positive and PFF-negative cells in increasing concentration of PFF 488. e) Gating strategy of CRISPRa screen showing top and bottom 15% of PFF intensity, while accounting for slight FSC-A trend.

**Supplemental figure 2:**
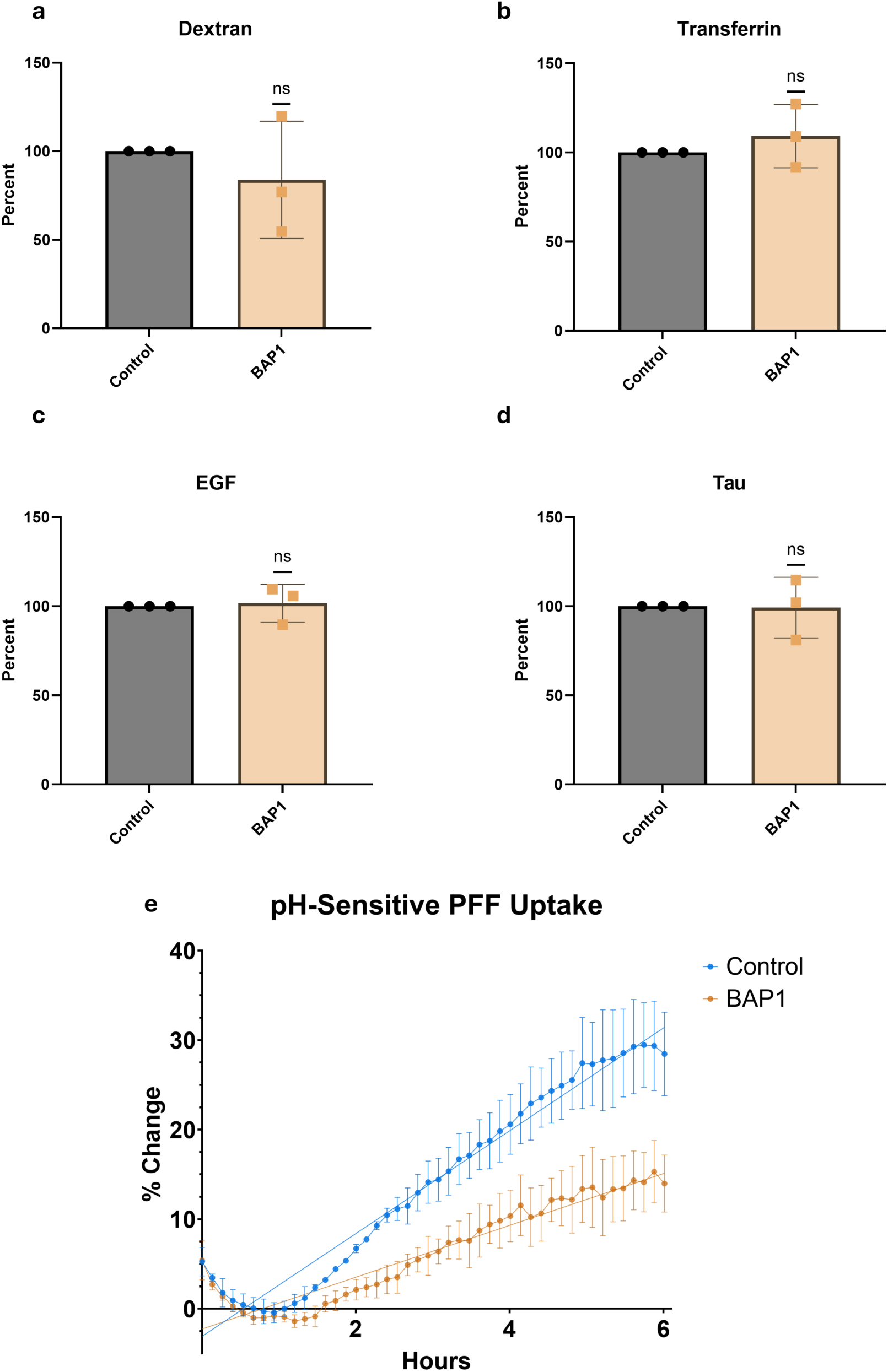
BAP1 does not affect cargo uptake and reduces lysosomal fibril internalization. a) Quantification of 24 hour 488-labelled 10k dextran uptake into CRISPRa control and BAP1a cells, normalized to control in each experiment. n=3 biological replicates. b) Quantification of 24 hour 488-labelled transferrin uptake into CRISPRa control and BAP1a cells, normalized to control in each experiment. n=3 biological replicates. c) Quantification of 24 hour 488-labelled EGF uptake into CRISPRa control and BAP1a cells, normalized to control in each experiment. n=3 biological replicates. d) Quantification of 24 hour TRITC-labelled tau oligomers uptake into CRISPRa control and BAP1a cells, normalized to control in each experiment. n=3 biological replicates. e) Time course of lysosomal internalization of pHrodo labelled PFF (pH-sensitive) in CRISPRa control and BAP1a cells. Linear regression slope comparison show significant difference between BAP1a and CRISPRa controls (p<0.0001). Statistical Analysis: a-d) one sample t test. E) simple linear regression. Graphs are presented as mean ± SD.

**Supplemental figure 3:**
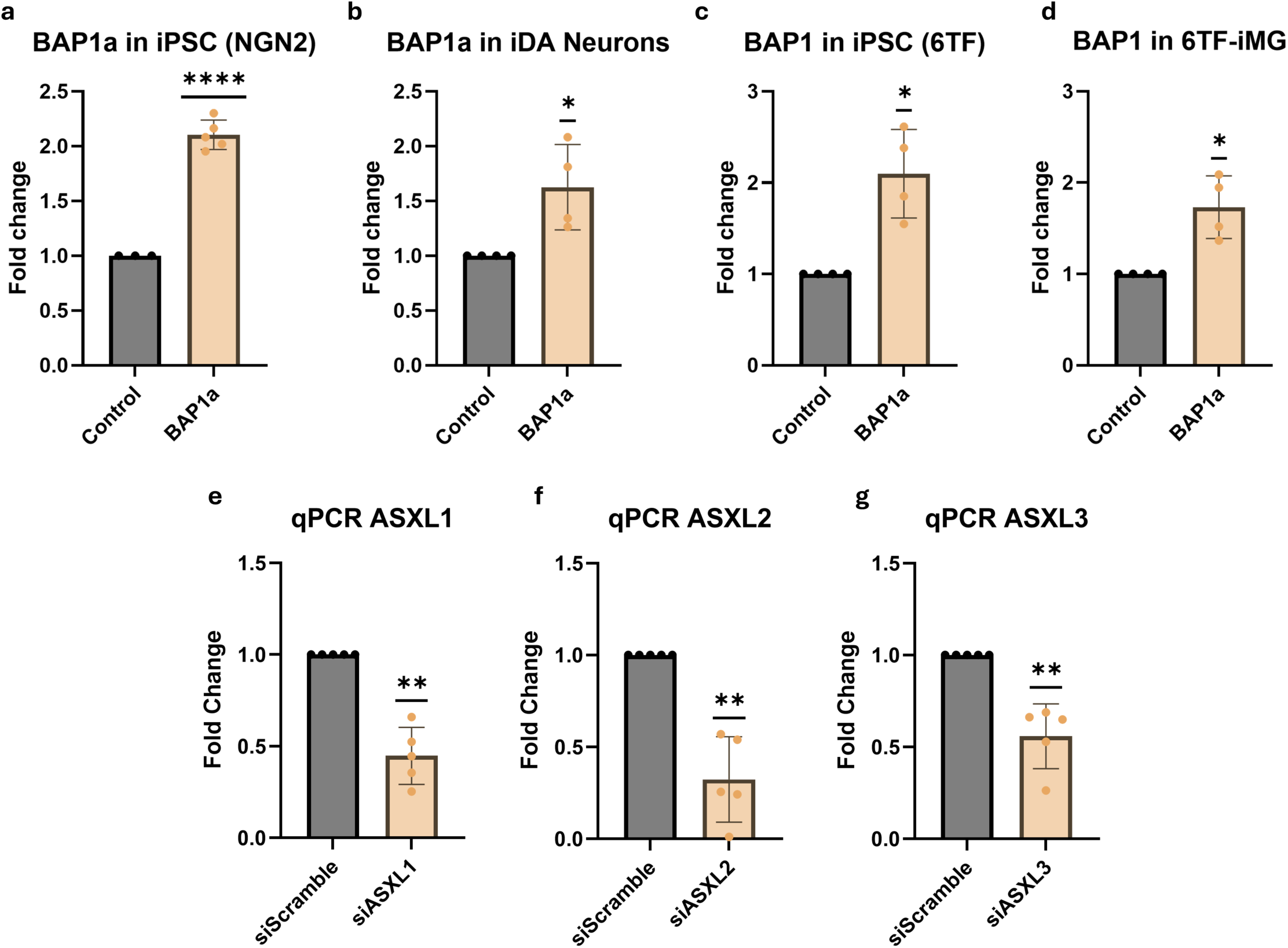
RT-qPCR vvalidation of CRISPRa and siRNA knockdown. a) RT-qPCR of *BAP1* CRISPRa activation in WTC11 iPSC compared to CRISPRa control, n=3-5 biological replicates. b) RT-qPCR of *BAP1* CRISPRa activation in WTC11 iDA compared to CRISPRa control, n=4 biological replicates. c) RT-qPCR of *BAP1* CRISPRa activation in 6TF WTC11 iPSC compared to sgRNA control control, n=4 biological replicates. d) RT-qPCR of *BAP1* CRISPRa activation in 6TF WTC11 6TF-iMG compared to sgRNA control control, n=4 biological replicates. e) RT-qPCR of *ASXL1* siRNA-mediated knockdown compared to siScramble, n=5 biological replicates. f) RT-qPCR of *ASXL2* siRNA-mediated knockdown compared to siScramble, n=5 biological replicates. g) RT-qPCR of *ASXL3* siRNA-mediated knockdown compared to siScramble, n=5 biological replicates. Statistical Analysis: one sample t test. Graphs are presented as mean ± SD. *p-value<0.05, **p-value<0.01, *** p-value<0.001, ****p-value<0.001

